# Effectiveness of yoga therapy as an adjunct on mental health status, quality of life, and medication adherence among people living with HIV on antiretroviral therapy: A randomized controlled trial (ART YOGA)

**DOI:** 10.1101/2025.08.26.25334511

**Authors:** D. Dhanlika, Partha Haldar, Neeraj Nischal, Naveet Wig, S. Shruti, S. Udisha, Rajesh Sagar, Bimal Kumar Das, Sanjay Ranjan, Gautam Sharma

## Abstract

**Introduction:** HIV, the retrovirus that causes AIDS, is a major global public health threat. This chronic viral infection diminishes the immune system by attacking CD4 cells. The principal treatment is antiretroviral medication (ART), which greatly increases the life expectancy of HIV patients. However, ART does not address psychological issues including depression, anxiety, and stress. These mental health issues are significant predictors of poor medication adherence, which can compromise treatment success. Depression and anxiety are associated with lower quality of life (QoL) and poor physical health because of diminished immunity. This trial aims to assess the effectiveness of yoga as an adjunct therapy on psychologic parameters (depression, anxiety and stress), quality of life, and medication adherence of people living with HIV on antiretroviral therapy at a tertiary care hospital in AIIMS New Delhi, India.

**Materials and methods:** This study is a two-arm, parallel-group, open-label blinded end-point trial, single-center, randomized controlled trial investigating the effects of Yoga Intervention as adjunct in People living with HIV (PLHIV). Participants (n=192) will be randomized to either 12 weeks of Yoga therapy program (n=96) or Active control group i.e., prescribed brisk walk (n=96). Both groups will receive standard treatment. The primary outcome is anxiety and depression scores (HADS-A and HADS-D), and the secondary outcomes are Stress (PSS), quality of life (WHOQOL-HIV BREF and SF-36 QoL) and medication adherence.

The study is approved by Institute Research Board Ethics (AIIMSA2969/03.01.2025, RP-46/25, OP-16/02.05.25) and is registered at Clinicaltrials.gov (CTRI/2025/03/081645). CTRI Link-https://www.ctri.nic.in/Clinicaltrials/pmaindet2.php?EncHid=MTIyNjUx&Enc=&userName=HIV,%20Yoga

## Introduction

Human immunodeficiency virus (HIV), the retrovirus responsible for acquired immunodeficiency syndrome (AIDS), remains a major global public health concern.[1] As of 2022, an estimated 39 million individuals were living with HIV worldwide, with 630,000 deaths attributed to AIDS-related causes.[2] In India, HIV prevalence among adults is approximates 0.22% (0.17–0.29%). [3]

HIV infection causes intricate innate and adaptive immunological reactions. Despite activation of CD4 T cells and CD8 T cell, these defense mechanisms are inadequate to eradicate the virus.[2,4] Currently, there is no definitive cure for HIV.[5] The effective ART/HAART (highly active antiretroviral therapy) is the cornerstone treatment for HIV infection, which acts on the suppression of viral replication, thereby preserving the immune function and prolonging survival thus transforming HIV as a chronic condition. [6–8] If left untreated, HIV may progress to acquired immunodeficiency syndrome (AIDS), marked by severe immune suppression and development of HIV-related malignancies or opportunistic infections.[9]

Observation studies indicate that ART medication adherence (below 90%) is significantly associated with lower odds of getting viral suppression.[10] Mental health issues such as depression and anxiety are the predictors of poor medication adherence to ART in PLHIV contributing to decreased quality of life.[11,12] These challenges arise from physical symptoms, emotional distress, societal stigma and social isolation, all of which impact mental and physical health partly due to immune dysregulation and decreased immunity [13–15] Depression and anxiety have shown to be associated with poor quality of life (QoL), which can impair physical health by lowering immunity. [16]

Both HIV infection and the use of HAART are associated with numerous immune-metabolic and morphological changes, such as chronic inflammation, lipodystrophy, dyslipidemia, diabetes, cardiovascular disease, and accelerated aging.[17–23] A study by Effendy E et al. reported moderate positive connection (r = 0.548) between CD4 levels and BDI scores highlighting the correlation between immune function and psychological well-being in people living with HIV(PLHV). [24] The complex interaction between the virus, treatment, and overall health, demanding comprehensive integrative management approaches for people living with HIV. Yoga is an age-old discipline that combines physical postures, breathing techniques, and meditation. Evidence suggests that even a short duration i.e., 10-days yoga-based lifestyle intervention significantly reduced IL-6 and TNF-α levels while increasing β-endorphin levels in patients with stress and inflammation.[25] Similarly an 8-week mindfulness meditation program improved CD4+ T lymphocyte counts in HIV-1-infected individuals.[26] Yoga also showed the improvement in clinical insight and medication adherence in schizophrenia patients.[27]

In PLHIV receiving combination ART, yoga has been associated with reductions in resting systolic and diastolic blood pressure compared to standard care.[28] Moreover, Yoga significantly improved functional capacity and health-related quality of life (HRQL) compared to standard care in HIV-seronegative population with heart disease, stroke, and COPD.[29] A recent meta-analysis found that yoga intervention significantly improved perceived stress (k=3), positive affect (k=3), and anxiety (k=2) compared to non-yoga control conditions in PLHIV. However, it reported no significant changes in depression and quality of life (k=2) possibly due to the small sample sizes and pilot nature of the included studies, as well as the lack of long-term follow-up data.[30] Given the current evidence and limitations of existing research, there remains a paucity of data supporting the effect of yoga-based interventions in PLHIV. There is a clear need for well-structured yoga-based intervention study that may help improve mental health status, quality of life and medication adherence of PLHIV.

### Rationale

PLHIV often experiences major psychological challenges such as anxiety, depression, and stress, which are worsened by the stigma, social isolation, and chronic nature of disease, impacting both their mental health, quality of life (QoL) and medication adherence. The need for more comprehensive and holistic therapies is highlighted by the fact that, although ART increases the life expectancy of people living with HIV by decreasing viral replication, it does not directly address psychological burden or quality of life. There is still a significant gap regarding yoga’s effect on PLHIV, despite its demonstrated advantages in enhancing psychological health and quality of life in a wide range of populations. Firstly, marked heterogeneity in the yoga forms (e.g., Hatha, Integrated, Sudarshan Kriya) and its durations in the existing studies, limit the understanding of which specific practices are most effective for improving psychological outcomes and quality of life (QOL). Secondly, study design (pilot trials) with small sample sizes and short follow-up periods, further result in inconsistent findings of the desired outcomes restricting its generalizability. This would be the first study aimed at investigating the effect of yoga interventions on medication adherence besides psychological outcomes and quality of life in PLHIV.

### Study Objective

The trial aims to assess the effectiveness of yoga as an adjunct therapy on psychologic parameters (depression, anxiety and stress), quality of life, and medication adherence of PLHIV on ART at a tertiary care hospital in AIIMS New Delhi, India.

### Research Questions

1. What is the effect of yoga as an adjunct therapy on Depression and anxiety scores among people living with HIV (PLHIV) on ART?
2. What is the effect of yoga as an adjunct therapy on Quality of Life among PLHIV on ART?
3. What is the effect of yoga as an adjunct therapy on Perceived Stress among PLHIV on ART?
4. What is the effect of yoga as an adjunct therapy on ART medication adherence among PLHIV on ART?

### Hypothesis

The primary hypothesis to be tested is 12 weeks yoga intervention as an adjunct therapy will improve depression and anxiety scores among PLHIV on ART therapy as compared to active control (moderate-to-intensity walk). The secondary hypothesis included 12 weeks yoga intervention will have positive effects on all the secondary health outcomes and 12-weeks yoga intervention will improve medication adherence in PLHIV on ART therapy as compared to active control (moderate-to-intense walk).

## Material and methods

### Trial design

We propose a two-arm parallel, prospective randomized, open-label blinded end-point trial, with four assessment points which include baseline, 3^rd^ month, 6^th^ month and 12^th^ month. The total study duration is three years which includes 1 year follow up.

### Study Setting

The trial will take place at the ART clinic, All India Institute of Medical Sciences (AIIMS), New Delhi for screening, enrollment, and randomization. The assessments, interventions, and follow-ups will be conducted at the Center for Integrative Medicine and Research, AIIMS New Delhi. The study protocol was created in compliance with the SPIRIT 2013 guidelines, the SPIRIT figure of the study illustrated in Figure 1, including the schedule of screening, enrolment and follow-up visits.[31] The study protocol is depicted in Figure 2. The visual abstract is presented in Figure 3.

**Figure 1:**
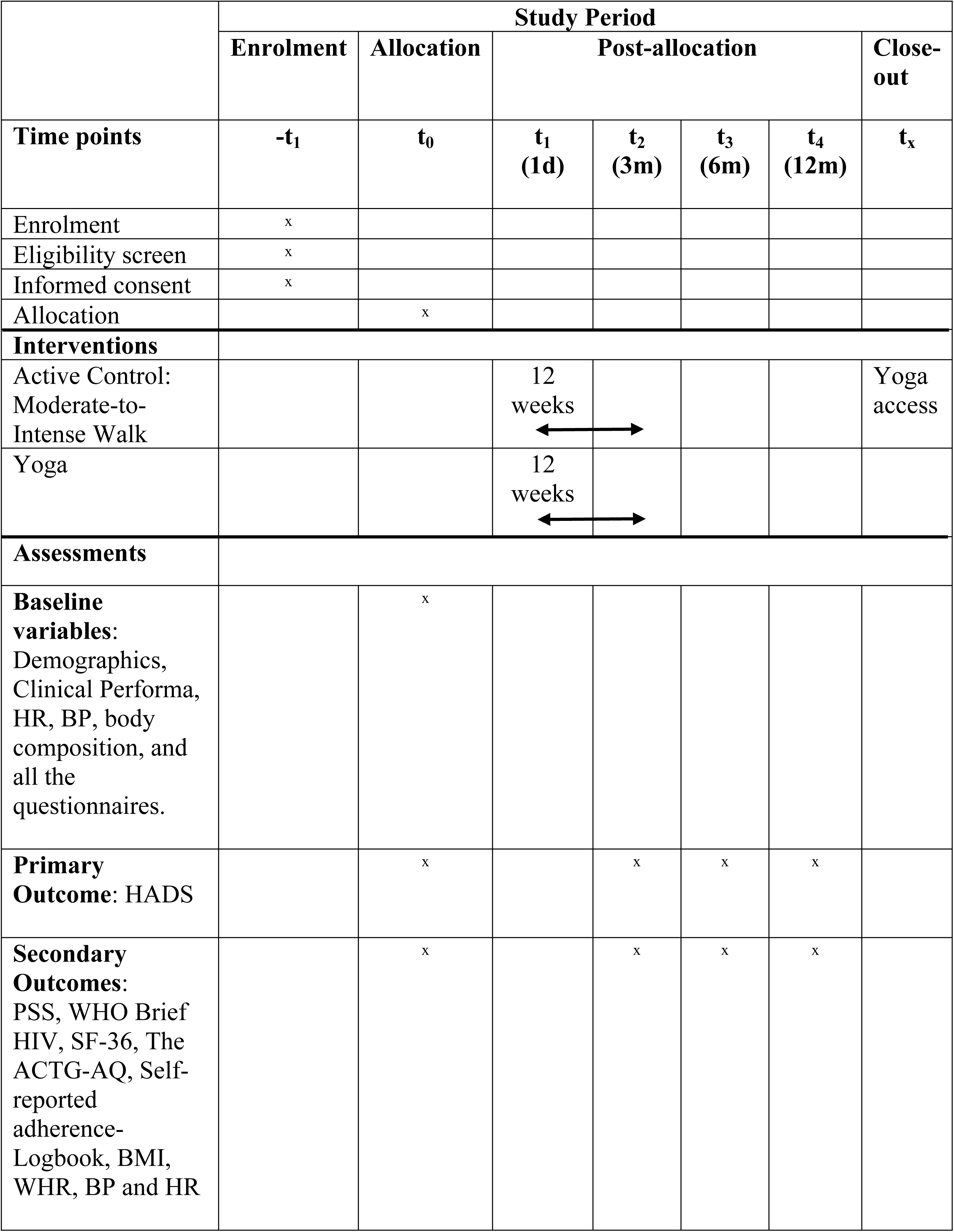
Based on Spirit guideline, Schedule of enrolment, Intervention, Baseline, Post-Intervention, Follow-up Assessment.^47^ HADS, Hospital anxiety depression scale; PSS, Perceived Stress Scale; WHO, World Health Organization Quality of Life; HIV BREF SF-36, 36-item Short Form Health Survey; The ACTG-AQ, The AIDS Clinical Trials Group Adherence Questionnaire (ACTG-AQ); BMI, Body Mass Index; BP, Blood Pressure; HR, Heart Rate; WHR, Waist-to-hip ratio.

**Figure 2.**
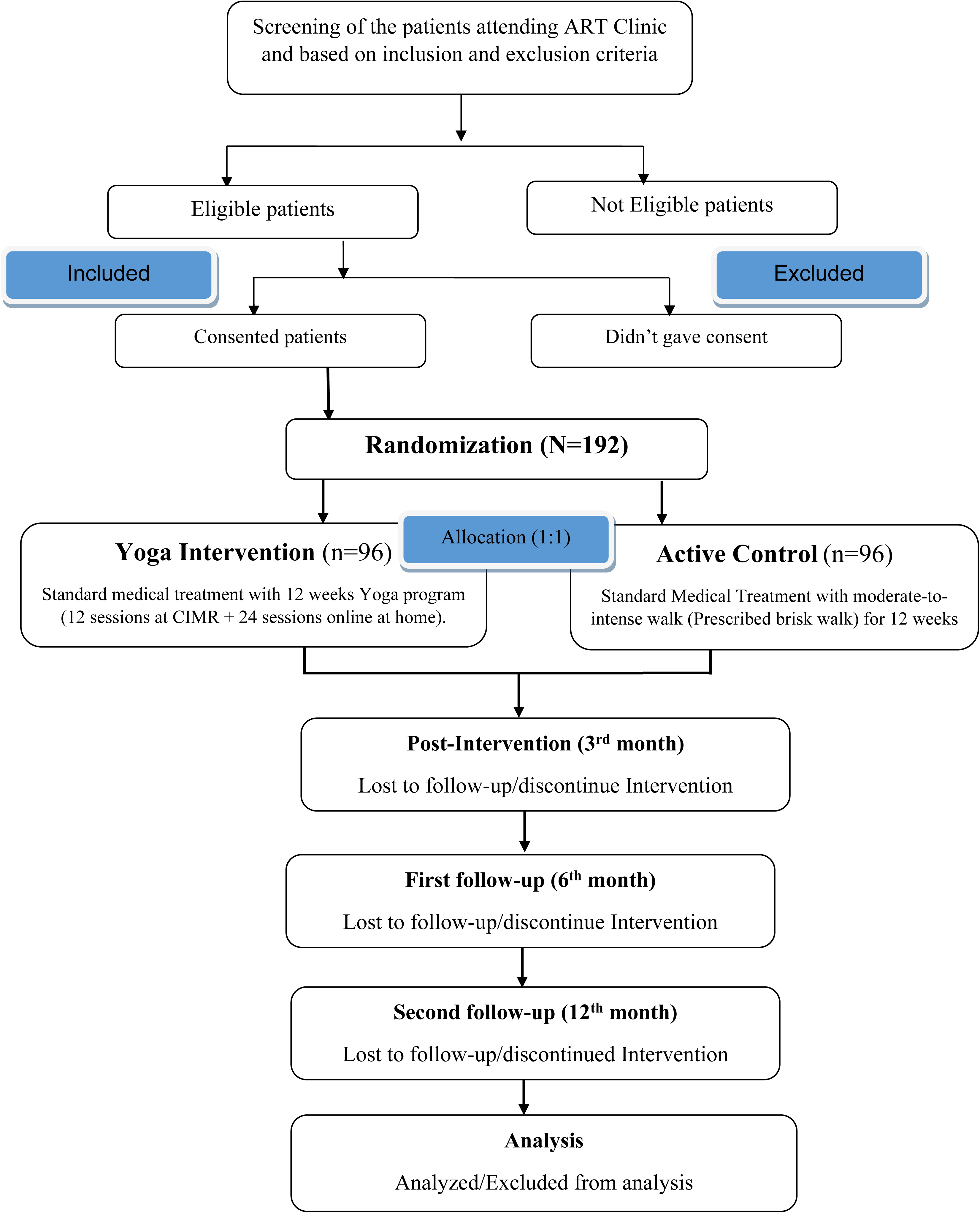
Study protocol.

### Participant Characteristics for the Clinical Trial

Individuals have been diagnosed as PLHIV as per the National AIDS Control Organization (NACO) guidelines. Eligible participants must have a most recent CD4 lymphocyte count of at least 350 cells/mm³, and HIV RNA viral load performed within 6 months below 50 copies/ mL (TND-Target Not Detected). A Karnofsky performance scale of more than 70 and be receiving standard antiretroviral therapy (ART) ≥ 6 months provided by the National AIDS Control Organization (NACO) or an equivalent recognized body. Additionally, they must undergo regular clinical follow-ups at the ART Clinic, AIIMS, New Delhi.

### Eligibility criteria

The subjects will be screened from ART Clinic, AIIMS New Delhi based on the following inclusion and exclusion criteria.

The inclusion criteria:

1. Diagnosed with HIV RNA, and on stable Antiretroviral therapy
2. Age: 18-50 years.
3. No contradictions to initiating a Yoga regime.
4. Karnofsky performance scale of more than 70.

Exclusion Criteria:

1. Current or history of hypertension, diabetes, heart failure, stroke and apparent opportunistic infection.
2. Use of antihypertensive drugs and anti-depressant medication.
3. Use of Immunosuppressive or immunomodulatory medications other than ART (within 6 months).
4. Pregnant and lactating women.
5. Known physical disability or mental disorder.
6. Practicing any form of yoga, meditation or exercise.

### Standard treatment

For the current trial, standard treatment is defined as the ART medication prescribed by the treating physician at ART clinic, AIIMS Delhi.

### The Yoga Intervention Arm

The yoga therapy program for the study is built on available evidence of yoga practices that demonstrate positive effects on the mental health and quality of life of PLHIV. An interdisciplinary team at CIMR conducted group discussion to design a yoga module, based on the current literature and the underlying pathophysiological mechanisms of the disease and its progression. The resulting module was then validated by 10 Yoga professionals representing different schools of yoga. The finalized yoga module included breathing exercises, loosening exercises, Asanas, Pranayama, and Relaxation Techniques. To ensure clarity, transparency, and reproducibility, yoga intervention will be recorded using TIDieR-Rehab (Template for Intervention Description and Replication) checklist (Table 1).[32]

**Table 1:** The TIDieR-Rehab (Template for Intervention Description and Replication for Rehabilitation) checklist.

| Item | Description |
| --- | --- |
| 1. Brief name | Yoga Intervention |
| 2. Why | Yoga has demonstrated benefits in improving psychological well-being and quality of life in the HIV seronegative population. However, there are several gaps in the current literature on yoga interventions for PLHIV. |
| 3. Who | People Living with HIV visit ART Clinic, AIIMS, Delhi |
| 4. When | Intervention will start within 7 days after Randomization for 12 weeks |
| 5A. What – Materials | For the yoga therapy sessions, chairs, bands, yoga mats, and wooden blocks will be used.<br>A PowerPoint presentation supported by a yoga manual for yoga therapists describing the intervention. Video of the complete yoga session and picture-based modules will be also given. |
| 5B. What – Procedures | Structured Validated Yoga Module (Table 3) |
| 6. Who provided | The yoga therapy program will be delivered by Yoga therapists, supported, and trained for delivering the intervention by the project team. Yoga therapists/Healthcare providers delivering the program will attend a specific one-day course to be certified to deliver the intervention program to ensure program fidelity. |
| 7. How | Both delivering face-to-face individually and online, using demonstration methods, study materials, and videos. |
| 8. Where | The intervention will be given at the C.I.M.R, convergence block, AIIMS, New Delhi. |
| 9A. How much – Session(s) duration/Frequency/ Intervention length | **Yoga Program:** The 12-week, 36-sessions program is divided into two parts.<br>a. 12 sessions for 4 weeks having 60-minute individually supervised yoga sessions with a yoga therapist to be done at C.I.M.R<br>b. 24 sessions for the remaining 8 weeks are 60-minute online therapy sessions.<br>The 12-week program will be supervised by the yoga therapist to ensure that the participants |
|  | correctly learn the practice of asanas, chanting, or meditation and prevent any possible injuries. The yoga module comprises exercises, loosening exercises, Asanas, Pranayama, and Relaxation Techniques. |
| 10. How challenging | The dosage (e.g. frequency, duration, intensity, level of severity) of yoga will be tailored to the participant's abilities throughout the program to support progression and continuous improvements in symptoms during the 12 weeks. Any changes made during the intervention (e.g., reducing intensity due to participant fatigue) will be recorded and reported. |
| 11. Regression/<br>Progression | The progression will be guided using the Borg scale of perceived exertion for yoga Practices.[33] |
| 12A. Personalisation –<br>Needs | Each participant in the yoga intervention will be assessed using a pre-intervention evaluation that included the Borg scale score. |
| 12B. Personalisation –<br>Preferences | Those with limited movement will be given the option of chair yoga/Strapes/wooden blocks. Instructor-led will be modified based participants physical and mental ability to understand and perform. The sessions are also designed with cultural and personal comfort levels in mind. |
| 13. Protocol deviations | The program will be tailored to the conditions, characteristics, preferences, progression, and improvements of the participant. Modifications will be made and reported based on the participant's condition or in case of disease progression during the trial (if any). |
| 14A. How well – Plan | **Compliance and adherence:** To be compliant with the yoga intervention, every participant must attend at least 27 out of the 36 sessions (75%) Adherence to Yoga Intervention will be evaluated using class attendance logs, self-reported home practice logs, and telephonic calls (every 15 days). Participants' partners will be invited to join the sessions to enhance adherence. |
| 14B. How well – Actual | Adherence and compliance will be recorded by session attendance and participant self-report |
| 15A. Harms – Plan | Throughout the study, each participant will be closely monitored for any adverse reactions that might result from the yoga intervention and for any |
|  | signs of disease progression. In the event of adverse reactions or disease progression, the treating physician will use clinical judgement and safety factors to determine whether a participant should remain in the study. Participants will receive proper medical attention and any required modifications to their yoga intervention in case of disease progression or adverse reactions. |
| 15B. Harms – Actual | To ensure the safety and wellbeing of every participant in the trial, any adverse occurrences will be recorded, submitted to the ethics committee, and examined. |

#### Yoga Program duration

The 12-week, 36-sessions program is divided into two parts. The Intervention schedule is detailed in Table 2. The 12-week program will be supervised by the yoga therapist to ensure that the participants correctly learn the practice of asanas, chanting, or meditation and prevent any possible injuries. The components of yoga module are detailed in Table 3. Although intervention will not be actively given after 12 weeks, patients will be encouraged to self-practice at home at least 3 times per week, and adherence to home practice will be monitored to assess sustained effects.

**Table 2:** Intervention Schedule.

|  |  |
| --- | --- |
| Total Contact Classes at CIMR | 12 contact sessions-3 sessions per week for 4 weeks |
| Online sessions | 24 online sessions -3 sessions per week for 8 weeks |
| Total supervised yoga sessions | 36 sessions (including both contact and online sessions) |
| 1. Subjects will be further guided through live video conferencing & pre-recorded videos.<br>2. During the non-contact period, the subjects will be encouraged to practice the yoga module 60 minutes/day at least 3 days a week<br>3. Picture-based guided Yoga booklets will be provided. |  |

**Table 3:**
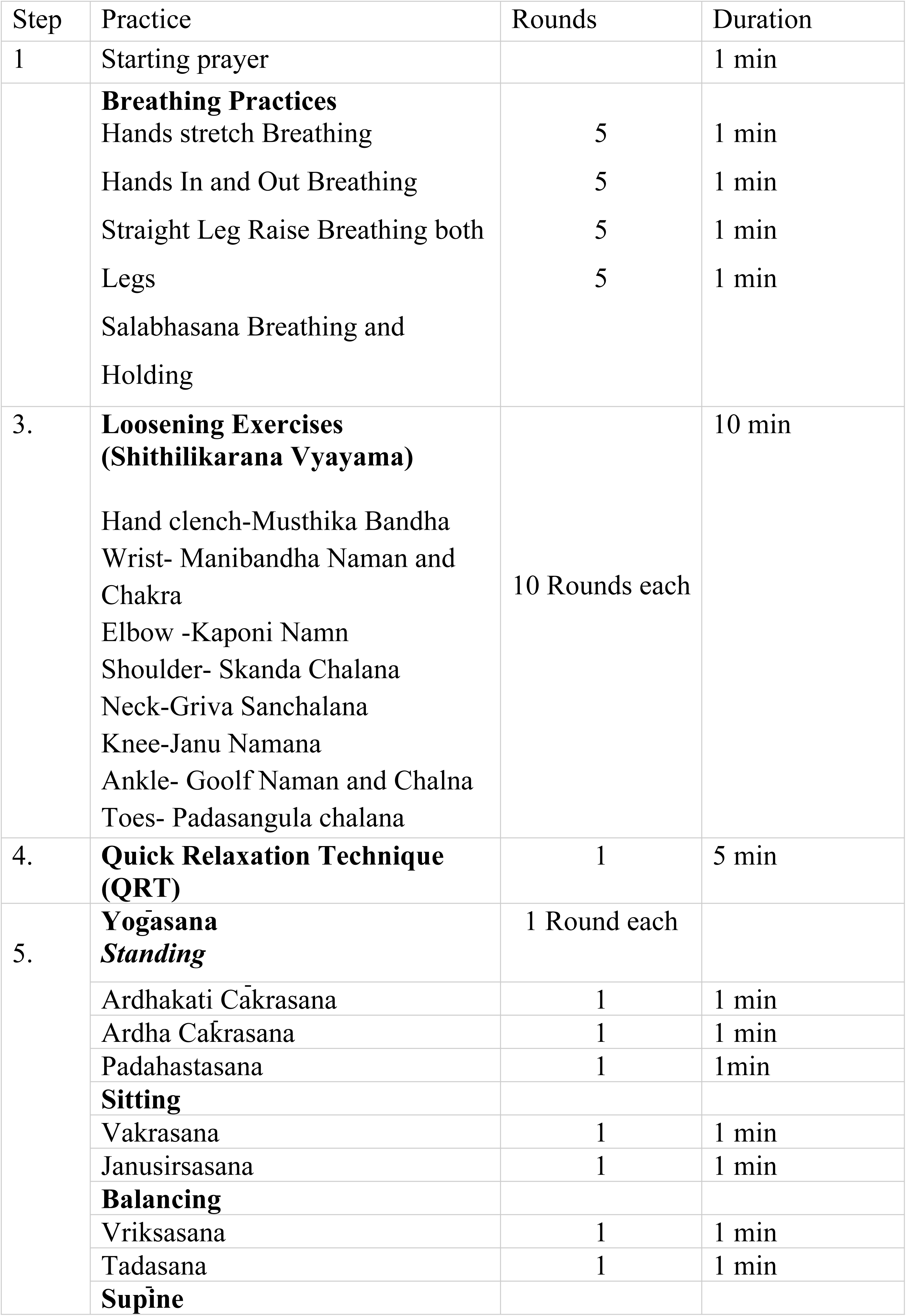
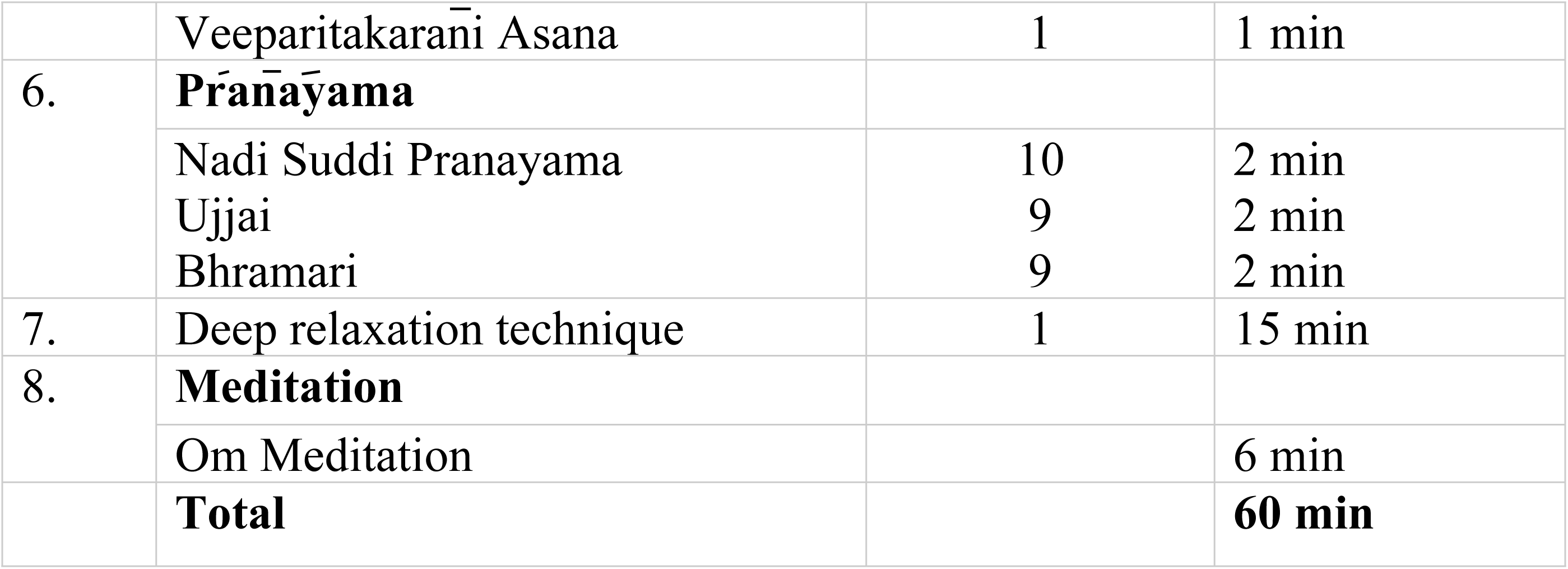
Integrated Yoga Module For PLHIV.

#### Compliance and adherence

To be compliant with the yoga intervention, every participant must attend at least 27 out of the 36 sessions (75%).

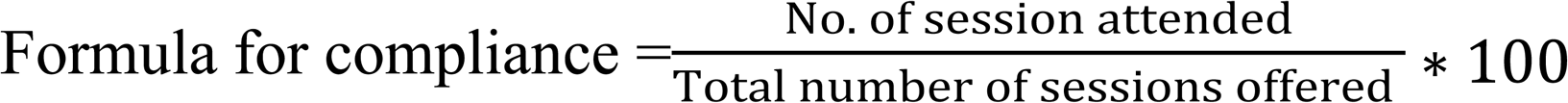

Adherence to Yoga Intervention will be evaluated using class attendance logs, self-reported home practice logs, and telephonic calls (every 15 days). Participant’s partners will be invited to join the sessions to enhance their adherence.

### Control group

The control group will be advised to continue the standard medical treatment for the same duration along with 12 weeks moderate-to-intensity walk per day (prescribed brisk walk). The duration of prescribed brisk walk will be at least practiced for a duration of 30 to 40 minutes every day, divided into three phases: a 5-minutes warm-up, 20-25 minutes of brisk walking, and followed by a 5–10 minutes cool-down. The provision of moderate-to-intensity walk in the control arm is to ensure the interest of the candidates to continue in the control arm, to reduce the attrition in the control group, and not limit them from potential risk reduction practices. Interested candidates from the control group will be taught yoga after the completion of the study. Both groups will be given a logbook to follow their sleep, diet, physical activities and intake of medications.

### Description of the Outcome measures

The study will begin with a baseline screening visit, at which written informed consent will be obtained before starting any additional study activity. The following information will be collected at baseline, 3^rd^, 6^th^ and 12^th^ month: clinical assessment, clinical history, demographic characteristics, and questionnaires. After being fully rested, the assessment will be done in the following order: Heart rate, blood pressure, body composition, and questionnaires (Hospital anxiety and depression scale (HADS), WHO QoL-HIV BREF, SF-36 questionnaire, Perceived Stress Scale and the ACTG Adherence Baseline & Follow-Up Questionnaire)

### Primary outcome measure

#### Hospital Anxiety and Depression Scale (HADS)

In 1983, Zigmond and Snaith created the widely used Hospital Anxiety and Depression Scale (HADS), a self-assessment tool, to identify whether patients in non-psychiatric hospitals or clinics, potentially or probably have anxiety disorders or depression. The scale is made up of 14 items divided into two subscales: the HADS-A (Anxiety Subscale), which has 7 items assessing anxiety symptoms, and the HADS-D (Depression Subscale), which has 7 items assessing depression symptoms.

Each item is assessed on a 4-point Likert scale, with total subscale values ranging from 0 to 21. The scale has cut-off points to classify the degree of symptoms: 0–7 is normal, 8–10 is borderline abnormal (mild), and 11–21 is abnormal (moderate to severe). Higher scores indicate greater levels of anxiety or depression.[34] This tool has strong internal consistency for both measures in HIV population, with a Cronbach’s alpha of 0.83 and 0.84 for the anxiety subscale and depression subscale respectively.[35]

### Secondary outcome measures

#### WHO QoL-HIV BREF

The WHOQOL-HIV BREF is a standardized questionnaire designed to assess the quality of life (QoL) in individuals living with HIV. The internal consistency of the questionnaire ranges from 0.65 to 0.83.[36] The questionnaire contains 31 items organized into six domains: physical health, psychological health, level of independence, social interactions, environment, and religious/personal beliefs. Each item employs a 5-point Likert scale to record respondents’ perspectives, with a score of one (1) indicating low and negative perceptions and a score of five (5) indicating high and positive perceptions, reflecting a higher quality of life (QoL). Negatively phrased items are reverse scored, ensuring that all responses fall between 1 and 5. Quality of life scores are divided into three categories: scores of 80 or more indicate great QoL, scores of 60 to 79 indicate good QoL, and scores below 60 indicate poor QoL. [37]

#### SF-36 Questionnaire

The questionnaire is standardized and scored using an algorithm. It evaluates the quality of life across multiple domains in varied populations. It determined health status by assessing physical functioning, physical role constraints, physiological discomfort, general health perceptions, energy/vitality, social functioning, emotional role limitations, and mental health distress. The scoring ranges from 0 to 100, and the patient responds to each question with one of five options: never, rarely, sometimes very often, or always. The internal consistency reliability coefficients for the questionnaire are higher than 0.70. [38]

#### Perceived Stress Scale (PSS)

The Perceived Stress Scale (PSS) is a psychological tool that assesses the degree to which people regard their lives as stressful. It was developed by Sheldon Cohen in 1983. It consists of 10 items that measure on a 5-point Likert scale (0 = Never, 4 = Very often), with higher scores indicating greater perceived stress. Some items are reverse scored to balance positive experiences. The total scores range from 0 to 40, with categories of low (0-13), moderate (14-26), and high (27-40) perceived stress. The Cronbach’s alpha for the total PSS-10 score is 0.73.[39,40]

#### Medication Adherence

Patient logbook will be given to all the participants to assess physical activity, sleep, exercise, diet, weight management, the medication adherence through pill counts (proportion of doses taken) and attrition rate. A mean medication adherence rate of less than 95% will be considered as suboptimal adherence.

Self-reported adherence: How many times have you missed at least one dosage of your HIV medicine in the last 30 days?” Participants provided a number ranging from 0 to thirty. Based on this response, the “single-item self-reported adherence” for the last month was calculated using the following method.[41]

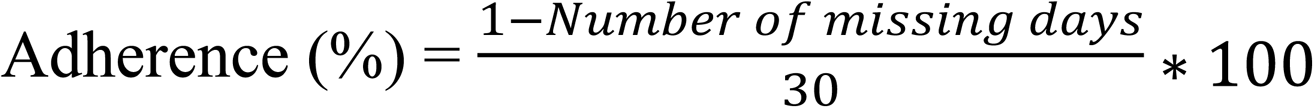

The AIDS Clinical Trials Group Adherence Questionnaire (ACTG-AQ)

It is a self-reported instrument that assesses HIV-positive person’s adherence to antiretroviral medication (ART). It was developed by the AIDS Clinical Trials Group (ACTG) and is intended to evaluate the degree to which individuals follow their recommended ART regimens for the past four days. Questions are related to adherence, non-adherence, and social factors associated with adherence. The questionnaire measures the patient’s response on the number of missed doses of a medication during each of the 4 days before a clinic visit. (E.g. How many doses did you miss yesterday, the day before yesterday, 3 days ago, and 4 days ago?). As patients usually take more than 1 drug, so that the adherence ratio for the previous four days is calculated using

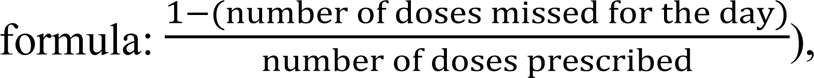

by doing so, we take the numbers of different drugs and the number of pills/doses into consideration The Cronbach’s alpha for the overall standardized scores (5 items/8 variables) is > 0.80. The scale is freely accessible for academic research use.[42]

#### Cardiac metabolic markers

Two consecutive resting blood pressure readings will be collected at 10-minute intervals while the participants are seated, with the cuff placed on the bare upper arm, one inch above the elbow. Blood pressure will be measured with calibrated, well-functioning digital blood pressure equipment (the Omron HEM 7124 Fully Automatic Digital Blood Pressure Monitor). Waist hip measurement: To measure the waist circumference, top of the hip bone and the bottom of the ribs will be located. Participants will be asked to exhale normally, and the tape will be placed halfway between these locations, aligned with their belly button. The tape will be wrapped around the waist, keeping it slacks enough to fit one finger inside. Finally, Participant will be asked to stand with their feet together and the measurements would be recorded. The measuring tape will be wrapped around the widest point of their hips and buttocks. Measurement closest to 0.1 cm is recorded.

Waist-hip ratio is measured using

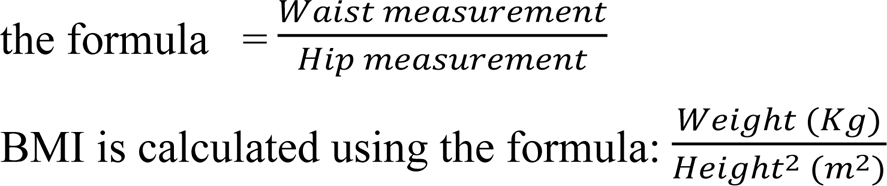

### Sample Size Calculation

The sample size for this study was determined by using R software to compare two means using a two-sample t-test. The calculation considered a pooled standard deviation (SD) of 3.38, a minimum important difference (MID) of 1.32 for HADS-Anxiety, effect size 0.39, a one-sided significance level (α) of 0.05, and 80% power (1 - β = 0.80). [43] The required sample size was determined to be 82 participants per group. To account for an expected 15% attrition rate over one year, the sample size increased to 96 participants per group. Thus,192 people will be recruited for the study (96 in each arm).

### Recruitment procedure

Patients will be screened from ART Clinic, AIIM, New Delhi. Eligible patients will be approached with a detailed patient information sheet explaining the study procedures, interventions, risks and benefits of participation, confidentiality, and expectations of participants. For eligible interested participants, informed consent will be obtained in writing, and they can withdraw consent at any time. A participant information sheet (PIS) in the local vernacular language will be provided to all eligible patients before randomization. The PIS document will detail the purpose of the study, measurements, procedures involved, and the risks and benefits of participation in the trial. (S1_Annexure1)

### Randomization

Once consented, and after baseline assessment, eligible participants will be randomized (1:1 allocation ratio) using permuted block randomization. A priori, a computer-generated randomization schedule prepared by an independent statistician in randomized, permuted block lengths. Variable block sizes (4, 6, or 8), to ensure group balance throughout the study and minimize predictability. Multiple small block sizes are chosen to limit the risk of anticipating the next group assignment while ensuring overall group balance throughout the trial. Allocation numbers will be concealed in opaque sealed envelopes, only accessible to a study staff. The envelopes will be opened after informed consent and baseline assessment. Blinding will be maintained for the outcome assessor and statistician analyzing the data, who will be unaware of group assignments. Baseline evaluations will be done by a nursing staff, and then the randomization will be done separate C.I.M.R staff who will not be part of assessment or intervention.

### Data collection and Data Management

EpiInfo, CDC’s free software, will be utilized for electronic data collection and management. It allows form customization, ensures standardized data entry, and provides offline data collecting when the internet is unavailable. Data entry forms in Epi Info will be pre-programmed with logic checks to prevent errors during data entering. All data gathering efforts shall follow Good Clinical Practice (GCP) principles. To maintain security and consistency, data will be coded in accordance with a predefined system. The Epi Info double-entry feature will be used to check data accuracy. Each data point will be entered twice, and any differences will be flagged for resolution via source document verification. An internal research study monitor will oversee the data gathering process to guarantee accuracy, integrity, and compliance with the study protocol. The monitor will periodically evaluate data entries and validate them against the source documents. Missing data will be handled using the imputation method of the last observation carried forward (LOCF). After the completion of the follow-up period of all the patients, data will be analyzed by an independent statistician, which will be blinded to the group allotment. A coded data will be handed over to the statistician with maintaining the blinding.

#### Criteria for dropout or early termination

##### Intervention Completion threshold

###### Dropout

will be considered if participants attend less than 27 sessions of a total 36 sessions being categorized as noncompliant and will be included in an intention-to-treat analysis (ITT). ITT analysis will include all participants as randomized, regardless of the number of sessions attended, any treatment switches, lost to follow-up due to disease progression (including development of opportunistic infection), or second-line treatment or death. However, such participants will be excluded from per protocol analysis. Per-protocol analysis will be limited to participants who meet the adherence threshold, i.e., attending at least 27 sessions.

##### Statistical analysis

The analysis will be performed using R software. The distribution of continuous data items will be examined using the Shapiro-Wilk test, and if needed the data distribution will be improved by performing log-transformation. The primary outcome, psychological status (HADS-Anxiety and HADS-Depression), will be assessed using a linear mixed model (LMM) to compare changes from baseline in HADS scores between the groups over time. Differences between groups at each time point will be reported as adjusted means with 95% confidence intervals (CIs). If the parametric analysis assumptions are invalidated, GEE models will be used. For quality of life (WHOQoL-HIV BREF and SF-36) and Perceived stress scale (PSS) will be analyzed using linear mixed-effects modeling, if required adjusting for baseline covariates. Medication adherence (ACTG adherence questionnaire and self-reported) will be evaluated using logistic regression. A multivariable logistic regression model will be used to identify independent predictors of adherence after accounting for demographic and clinical factors. An intention-to-treat analysis will be performed with a separate per-protocol analysis. A p-value of <0.05 will be regarded statistically significant.

###### Sensitivity Analysis

**-**Subgroup analysis will be done among participants who progressed from first line to second-line treatment or disease progression (including development of opportunistic infection), length of duration of disease (less than / more than 2 years) and those with low vs high adherence to evaluate how these factors impact the outcomes or adherence. Multivariable regression analysis will be used to investigate the associations between different secondary outcomes and the intervention, while adjusting for confounding variables such as disease stage, treatment adherence, and demographic features.

### Adverse Reactions Monitoring

Throughout the study, each participant will be closely monitored for any adverse reactions that might result from the yoga intervention and for any signs of disease progression. In the event of adverse reactions or disease progression, the treating physician will use clinical judgement and safety factors to determine whether a participant should remain in the study. Participants will receive proper medical attention and any required modifications to their yoga intervention in case of disease progression or adverse reactions. To ensure the safety and wellbeing of every participant in the trial, any adverse occurrences will be recorded, submitted to the ethics committee, and examined.

## Discussion

This randomized controlled trial aims to evaluate the effectiveness of yoga as an adjunct therapy on psychologic parameters (depression, anxiety and stress), quality of life, and medication adherence of PLHIV on ART at a tertiary care hospital in AIIMS New Delhi, India. While antiretroviral medication has made HIV a manageable chronic disease by reducing viral replication, preserving immune function, and extending life, [6–8] it does not address the mental health issues and poor adherence that frequently accompany the condition. Depression and anxiety, which are prevalent in PLHIV, have been demonstrated to affect physical health by reducing immunity and are predictors of poor QoL and inadequate ART adherence. [16] These challenges thus raise the possibility of virologic failure and diminish the effectiveness of ART. Yoga has been studied as a complementary therapy for improving mental health and QoL across diverse populations, including PLHIV. A recent meta-analysis reported significant improvements in perceived stress, positive affect, and anxiety among PLHIV undergoing yoga interventions.[30] However, no significant changes were observed in depression or QoL, likely due to methodological limitations of the included studies, such as small sample sizes and the absence of long-term follow-up. These limitations emphasize the critical need for well-designed, adequately powered studies to access the long-term impacts of yoga on this population’s mental health, quality of life, and adherence to ART. Our study is designed to address these gaps by implementing a well-structured, validated, 12-week yoga program that incorporates both psychological and behavioral outcomes, with rigorous methodology to generate high-quality evidence on the role of yoga as a complementary therapy in HIV care. The only limitation of the study design is that the outcomes like depression, anxiety, stress and quality of life depend on self-reported measures, which can introduce recall and response biases. The strength of the study is the yoga intervention in addition to ART focuses on the psychological burden, quality of life and medication adherence. The study focuses on the pleiotropic effect of yoga on medication adherence, an often overlooked yet crucial factor in long-term HIV management. Longer follow-up periods will allow for better observation of both short-term and sustained effects of Yoga Intervention. TIDieR-Rehab (Template for Intervention Description and Replication for Rehabilitation) checklist is used to provide extensive guidance on recording yoga interventions, emphasizing clarity, transparency, and reproducibility. If proven effective, the findings of the trial could drive a significant shift from managing PLHIV health to adopting a holistic approach that addresses the overall well-being, mental health, and quality of life of People Living with HIV (PLHIV) through yoga intervention, there might be an indication to encourage yoga practices systematically in clinics.

### Ethics and dissemination

the study is approved by Institute Research Board Ethics (AIIMSA2969/03.01.2025, RP-46/25, OP-16/02.05.25) and is registered at Clinicaltrials.gov (CTRI/2025/03/081645). All study findings will be published in scientific peer-reviewed journals and presented at scientific conferences.

### Protocol amendments

Any changes to the study protocol will be evaluated and approved by the Institute Research Board Ethics prior to implementation. All pertinent parties, including the Institutional Ethics Committee, CTRI registry, investigators, and, where appropriate, will be promptly informed of any significant protocol amendments. Versions of the updated protocol will be documented and submitted for ethics approval.

### Trial Status

Recruitment is planned to begin in November 2025, and data collection is estimated to be completed by December 2027.

## Data Availability

No datasets were generated or analysed during the current study. All relevant data from this study will be made available upon study completion (study protocol). Deidentified research data will be made publicly available when the study is completed and published

## Funding Status declaration

The study has been submitted for funding to the Indian council of medical research (ICMR) for “Investigator-Initiated research proposals for intermediate extramural grants (2025)” (8.4.2025). The outcome is currently pending at the time of manuscript submission.

## Contributors

DD conceived the study and led the development of the protocol. DD prepared the first draft of the manuscript and is the main methodologist. [DD] [PH] [GS] [NN] and [NW] contributed to the design of the intervention, selection of outcome measures, and refinement of the protocol. [DD] [PH] and [US] provided statistical guidance and drafted the statistical analysis plan. [RS] [NN] [NW] [BD]and [SR] contributed clinical expertise and assisted in revising the protocol to meet institutional and ethical standards. All authors reviewed and approved the final manuscript. [GS] is the guarantor of the study.

### S1_Annexure

Participant Information Sheet (PIS) and Participant Informed Consent Form (PICF).

### S2_SPIRITchecklist

SPIRIT 2013 Checklist.

---

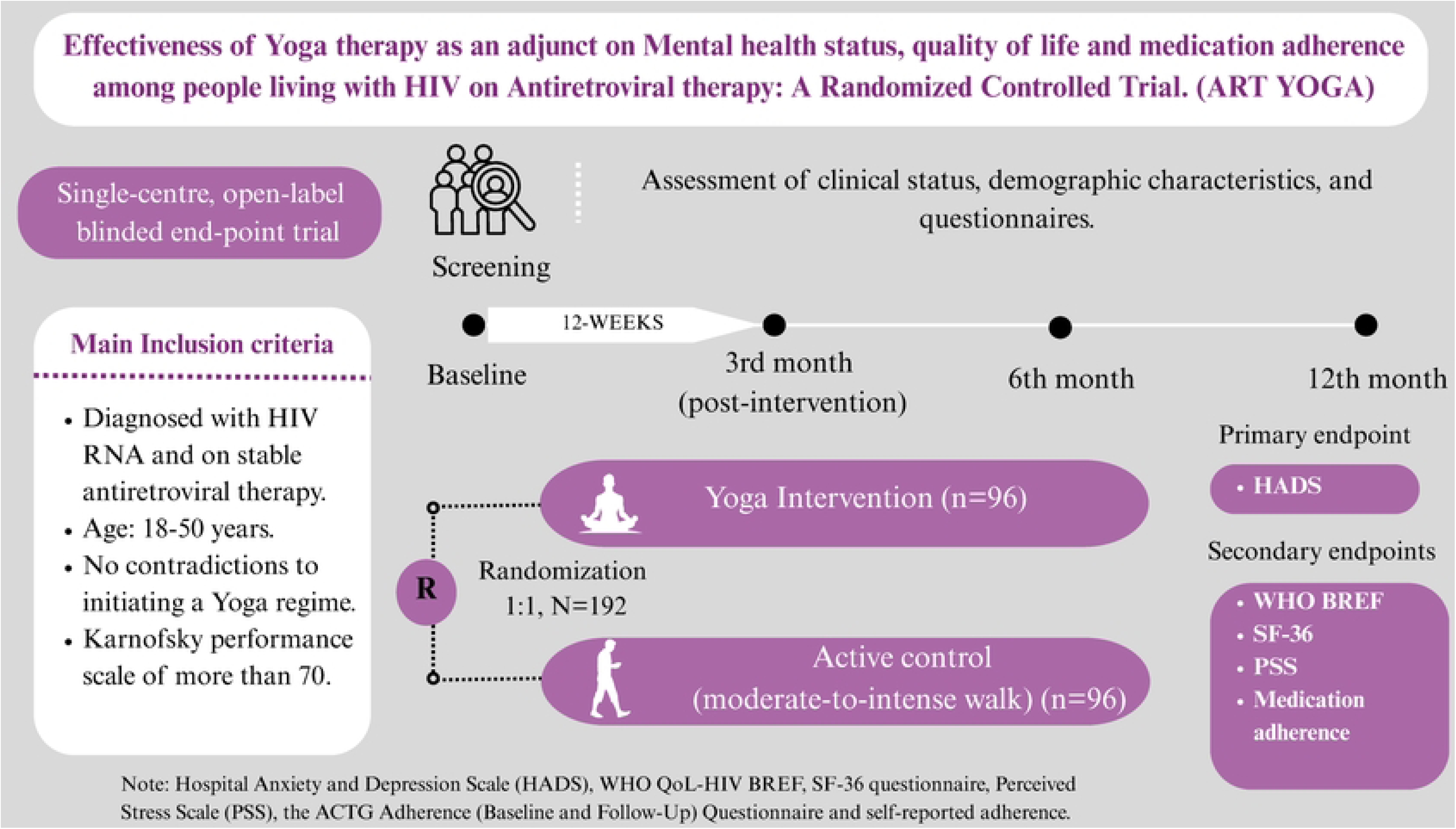

